# Patients Report Better Outcomes Following Reduction of Tibial Plateau Fractures: A Systematic Review

**DOI:** 10.1101/2025.01.08.25320009

**Authors:** David S. Kitchen, Sebastian Ricci, John M. Abrahams, Michael L. Downie, Gerald J. Atkins, Lucian B. Solomon, Peter J. Smitham

## Abstract

**Background:** Tibial plateau fractures (TPF) are complex injuries involving the articular surface of the proximal tibia, commonly managed with open reduction and internal fixation (ORIF). The aim of this management is to restore joint alignment whilst respecting the soft tissue envelope. The relationship between radiological factors on patient-reported outcomes (PROMs) remains unclear. This systematic review aimed to determine the association between articular reduction and PROMs following TPF.

**Methods:** MEDLINE, Embase, Cochrane CENTRAL, ClinicalTrials.gov and Google Scholar databases were searched for English language articles between January 2000 to 5 May 2023, with 3300 studies screened. Eligible studies reported radiological and clinical outcomes in adult TPF patients managed with ORIF, with a minimum follow-up of two years. Data extraction and quality assessment were conducted independently by two reviewers using the Risk of Bias Assessment tool for Non-Randomised Studies (RoBANS). Linear regression models assessed the effect of articular reduction on PROMs.

**Results:** A total of 30 studies met the inclusion criteria, comprising 1591 patients. Studies with >85% of cases achieving articular reduction within 2 mm reported significantly higher PROMs, particularly Hospital for Special Surgery (HSS) scores (p = 0.04). To analyse outcomes between studies that used different PROMs, a grouped analysis approach was utilised, which showed ‘excellent’ outcomes associated with cohorts in which higher rates of successful reduction were achieved when compared to ‘good’ outcomes (p = 0.04).

**Conclusions:** Achieving higher rates of articular reduction increases the likelihood of excellent patient-reported outcomes. Additionally, studies in which a larger proportion of the cohort achieved reduction reported significantly higher PROMs compared to those with fewer patients achieving anatomical reduction. This study also introduces a novel method to standardise the reporting of different PROMs in a meta-analysis, enabling comparison of heterogeneous data.

## Introduction

Tibial plateau fractures (TPF) are injuries of the articular surface of the proximal tibia that are commonly managed by open reduction and internal fixation (ORIF). The injuries follow a bimodal distribution with high-energy mechanisms in the younger and middle-aged cohort with a lower energy mechanism being more prevalent in the elderly age group (1).

Fractures of the tibial plateau often have marked depression and comminution of the articular surface, as well as significant soft tissue injuries. Surgical treatment is a balance between restoration of articular congruency and alignment whilst minimising additional soft tissue damage to avoid short and long-term complications with the goal of restoring function and reducing the risk of developing secondary osteoarthritis. This has resulted in a number of different strategies regarding management timing, surgical approaches and fixation techniques (2–5), including open reduction supplemented with arthroscopic-assisted techniques (6, 7).

Without restoration of the articular surface, the resultant joint incongruence may cause uneven loading and lead to joint degeneration (8). Despite surgical intervention there still remains a significant association with post-traumatic arthritis in TPF patients (9), with these same patients having an increased likelihood of undergoing total joint arthroplasty within 10 years of their injury (10, 11).

Restoration of the articular surface after TPF has long been the primary goal of surgical intervention (12). The need for anatomical articular surface reduction has been questioned, with several recent studies unable to find a strong association between restoration of the joint surface (within 2 mm) and patient reported outcome measures (PROMs) (13–17). Moreover, mechanical coronal malalignment and instability are thought to be important to patient outcomes (1, 18, 19). This has been associated with poorer outcomes in those with increased varus malalignment, with valgus alignment having a lesser influence on outcome (13, 16).

The purpose of this systematic review was to assess the relationship between radiological factors and PROMs following TPF. The secondary aim of the study was to determine whether restoration of the articular surface to within 2 mm impacted PROMs. To our knowledge, a review of the evidence comparing radiological, and patient reported outcomes in TPF has not been previously performed.

## Methods

### Search strategy

The systematic review was performed in accordance with PRISMA guidelines. A single search strategy, customised for each database, was developed using a mix of controlled vocabulary and keywords. Search results were reviewed to ensure identification of relevant studies. The search was limited to articles in English and published since 2000. Conference abstracts and animal studies were excluded. The following databases were searched until 5 May 2023: MEDLINE (Ovid), Embase (Ovid), Cochrane CENTRAL (Cochrane Library), ClinicalTrials.gov and Google Scholar (20). The full Embase search strategy is shown in Appendix A.

### Eligibility criteria

Inclusion criteria for the study were adult tibial plateau fractures with an articular component, Schatzker types I-VI or AO/OTA classification 41 type B or type C, who underwent primary operative intervention with internal fixation. Fracture reduction had to be reported with a mean follow-up time of at least two years. Radiological alignment was reported as either fracture reduction or coronal alignment. Clinical outcome was recoded as either a functional score or PROMs. Included studies were published in English between the years 2000-2023.

Exclusion criteria included reports on individuals under 18 years old and studies reporting on fractures of a pathological nature or associated with joint arthroplasty. Also excluded were studies solely addressing open fractures, and those involving additional fractures to the ipsilateral limb or vascular injuries. Published case reports, or studies in which definitive management included either external fixation or non-operative measures were excluded. Additionally, systematic reviews, abstracts-only, protocols, or comment articles without datasets, along with single-patient case reports, were excluded,

### Study Selection

Independent screening was performed by two authors (DSK and SR) by title and abstract, then by full text, as necessary based on inclusion and exclusion criteria. If consensus was not achieved these were resolved in discussion with a third author (PJS).

### Quality assessment

The included studies were independently assessed for risk of bias by two authors (DSK and SR). The studies were assessed using the Risk of Bias Assessment tool for Non-randomised Studies (RoBANS) (21). This tool assesses bias across several domains, including selection of participants, confounding variables, measurement of exposure, blinding of outcome assessment, incomplete outcome data and selective reporting. Each of these categories is then rated as ‘high’, ‘low’ or ‘unclear’. If consensus was not achieved these were resolved with discussion with a third author (PJS).

### Extraction of data

Two authors (DSK and SR) independently extracted data from the relevant studies into a standardised proforma. Consensus on data was achieved through discussion prior to statistical analysis.

### Statistical analysis

All statistical analysis was performed using GraphPad Prism 9.0 (GraphPad Software Inc, Boston, MA, USA). To assess whether articular reduction influenced PROMs, a linear regression analysis was performed for all studies that reported on the mean PROM and whether articular reduction was achieved. A separate liner regression analysis was performed based on the PROM (KOOS pain, KOOS QOL, Lysholm and HSS). The mean number of cases that had articular surface reduced was then used to categorise cohorts to determine if there was a significant difference in PROM values. Given the heterogeneity of PROMs scores used, further grouping analysis was performed categorising the respective outcomes into ‘excellent’ and ‘good’. Significance was determined using a Student’s t-test and was defined as *p* < 0.05.

## Results

The initial search retrieved 5,228 studies, which on removal of duplicates resulted in 3,300 unique studies to screen (**Fig. 1**). 224 full-text studies were then assessed, with 30 studies (**Table 1)** satisfying the inclusion and exclusion criteria, for a total of 1,591 patients assessed.

**Figure 1.**
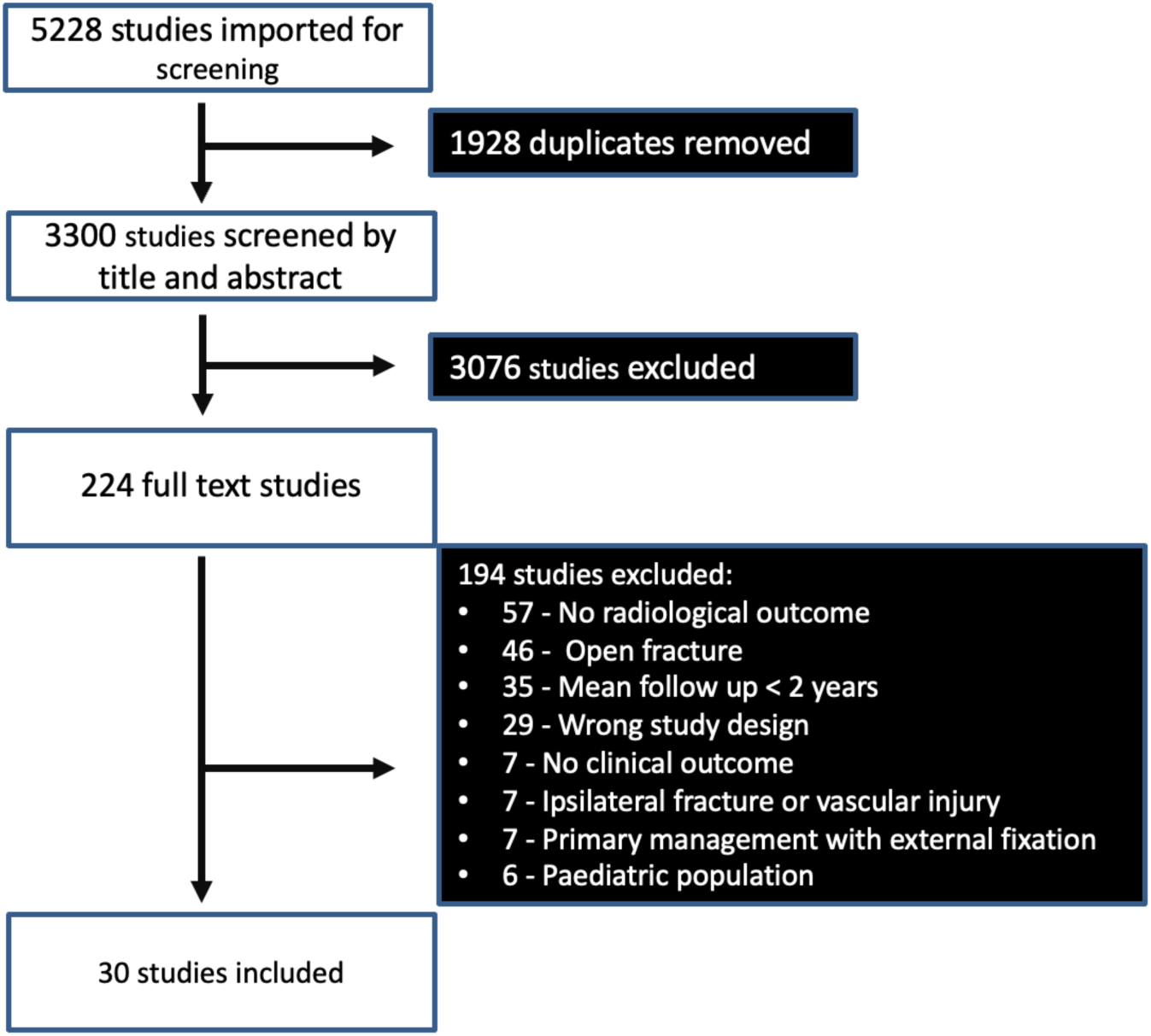
PRISMA flow diagram showing selection of studies for the systematic review.

**Table 1.**
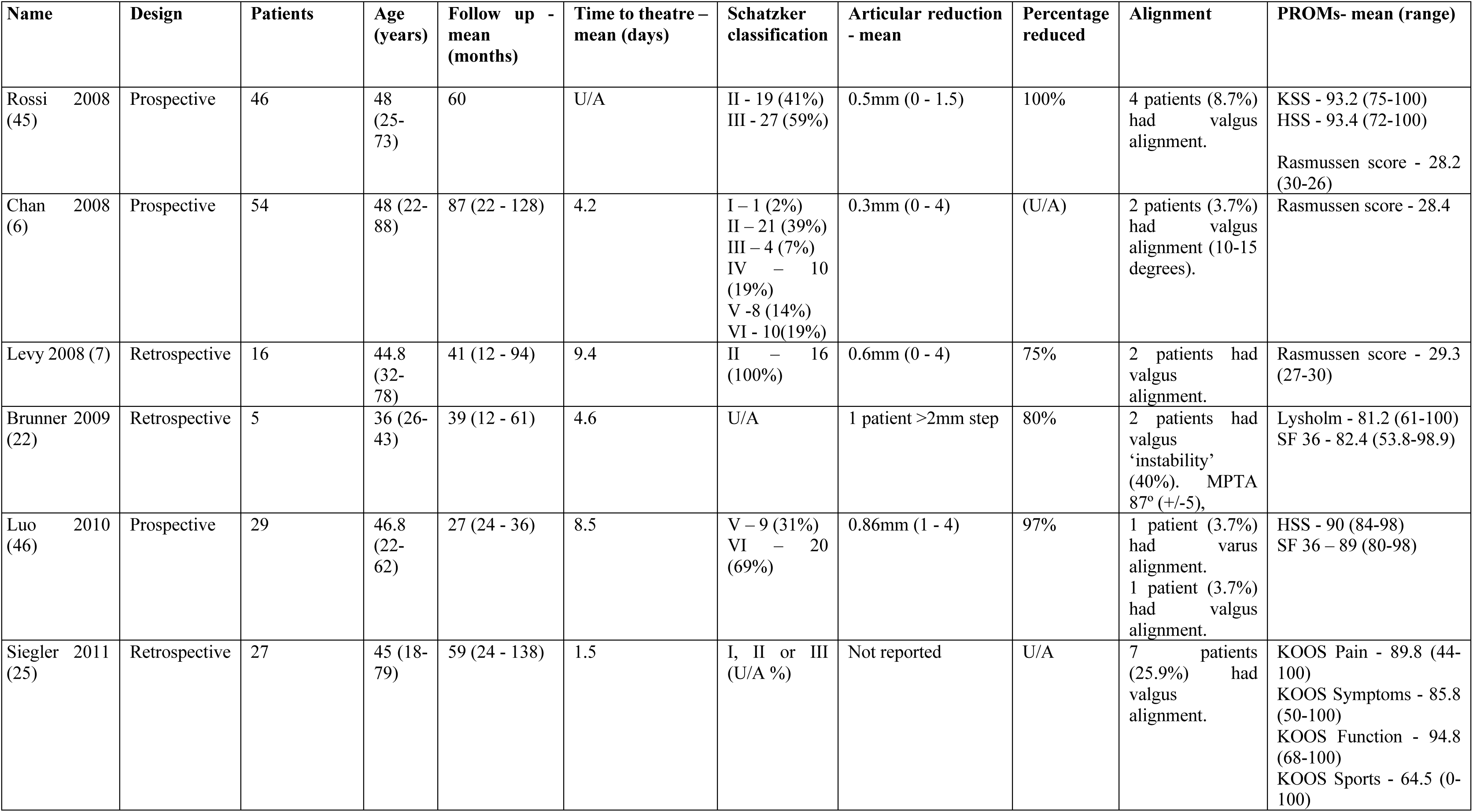
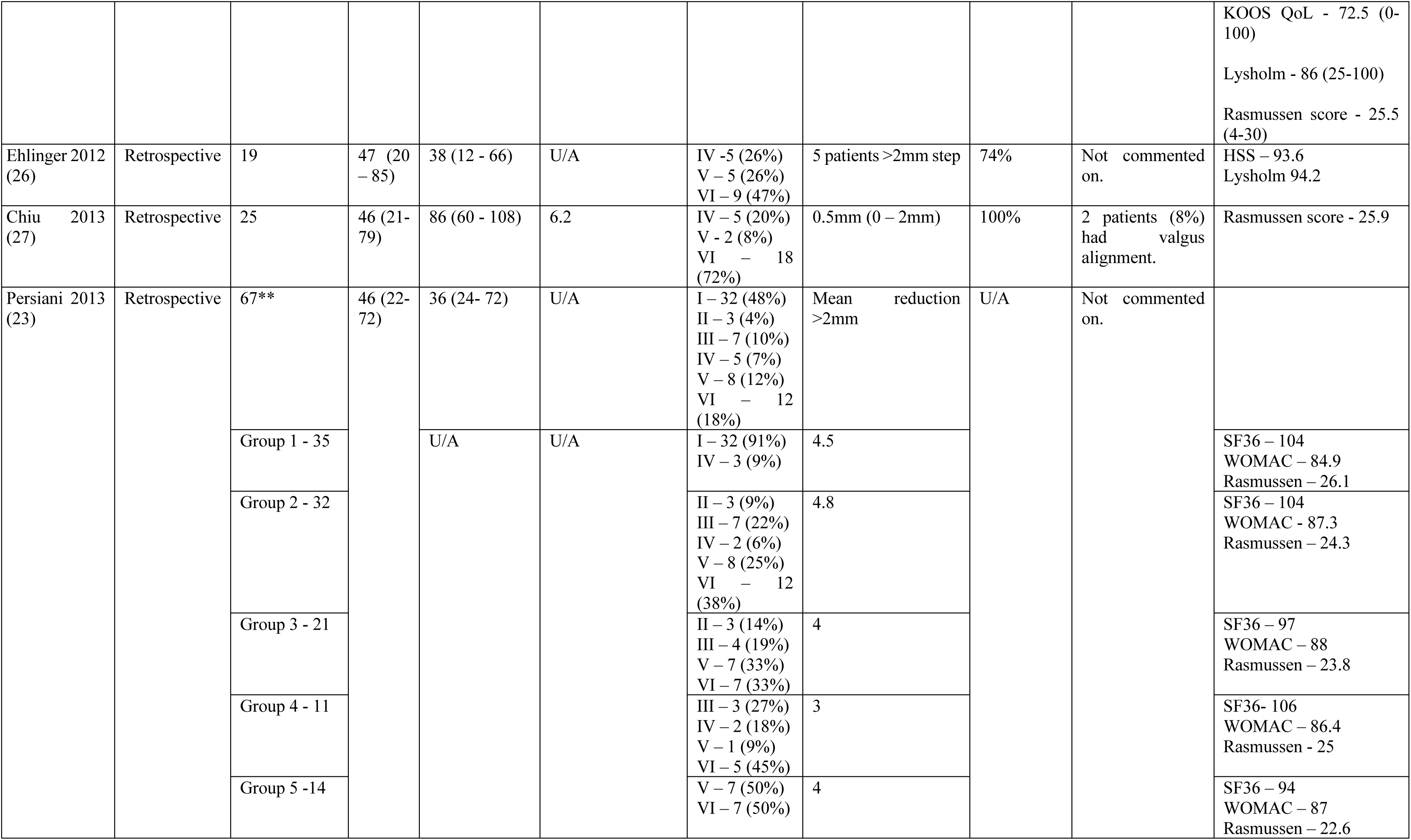
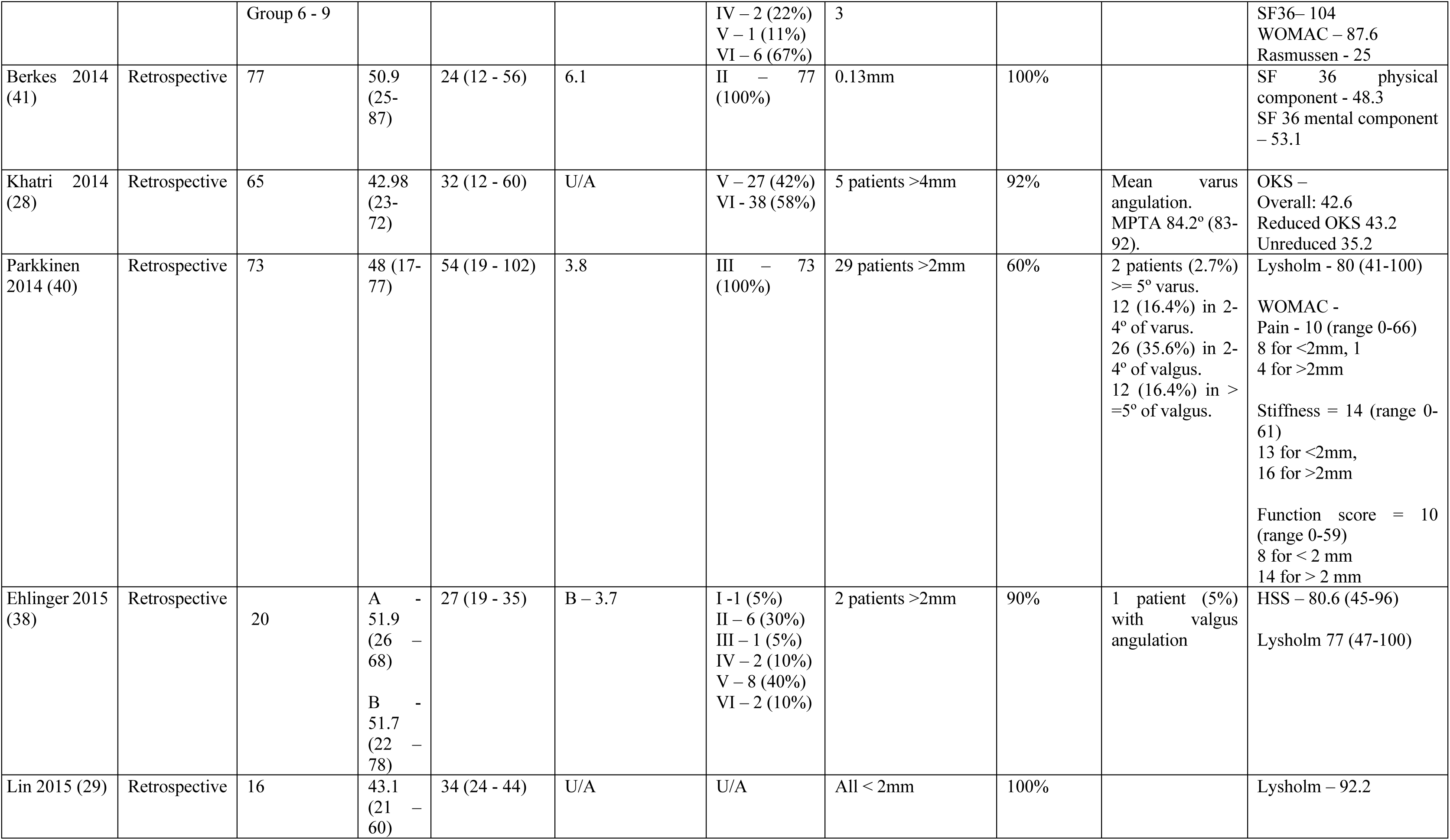
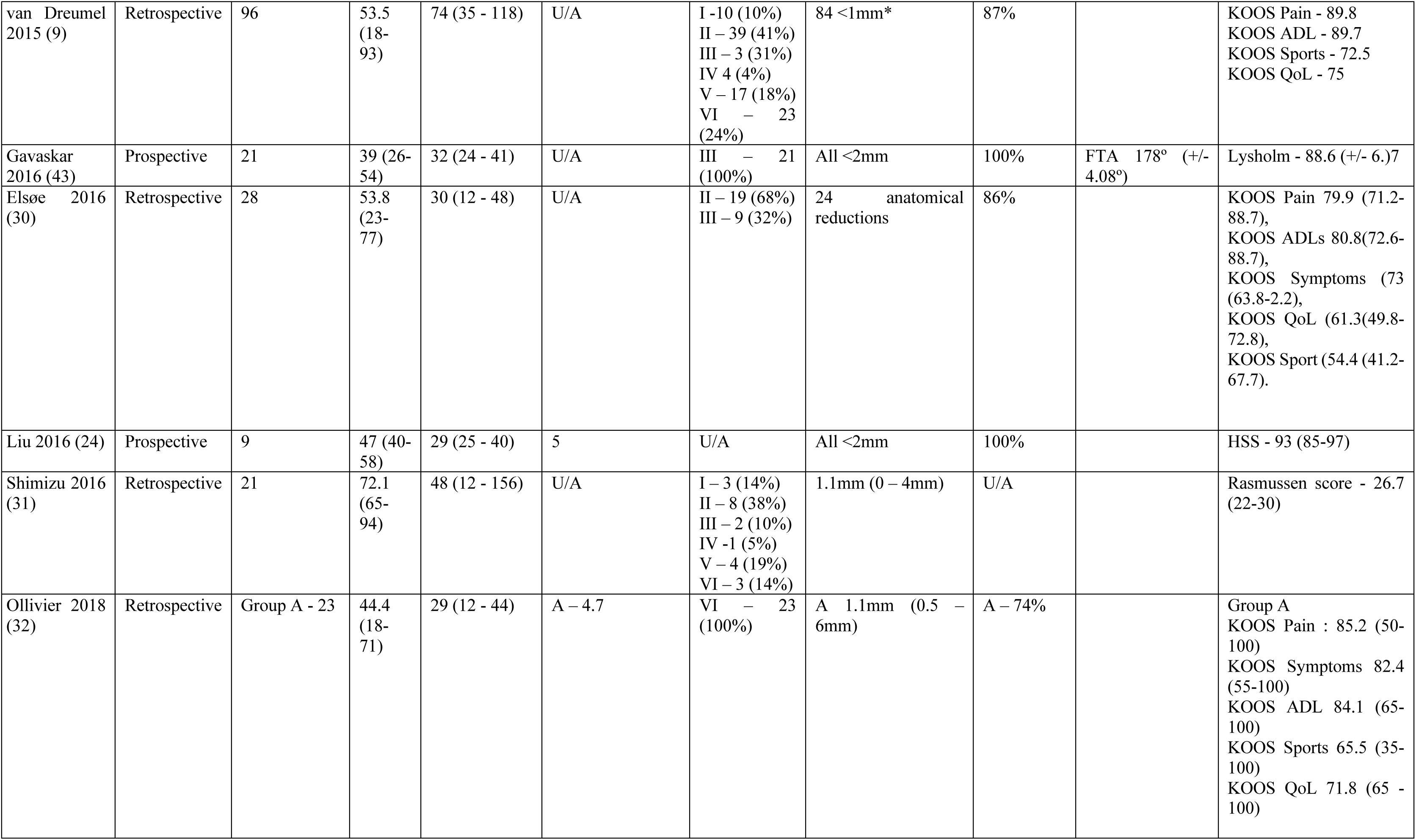
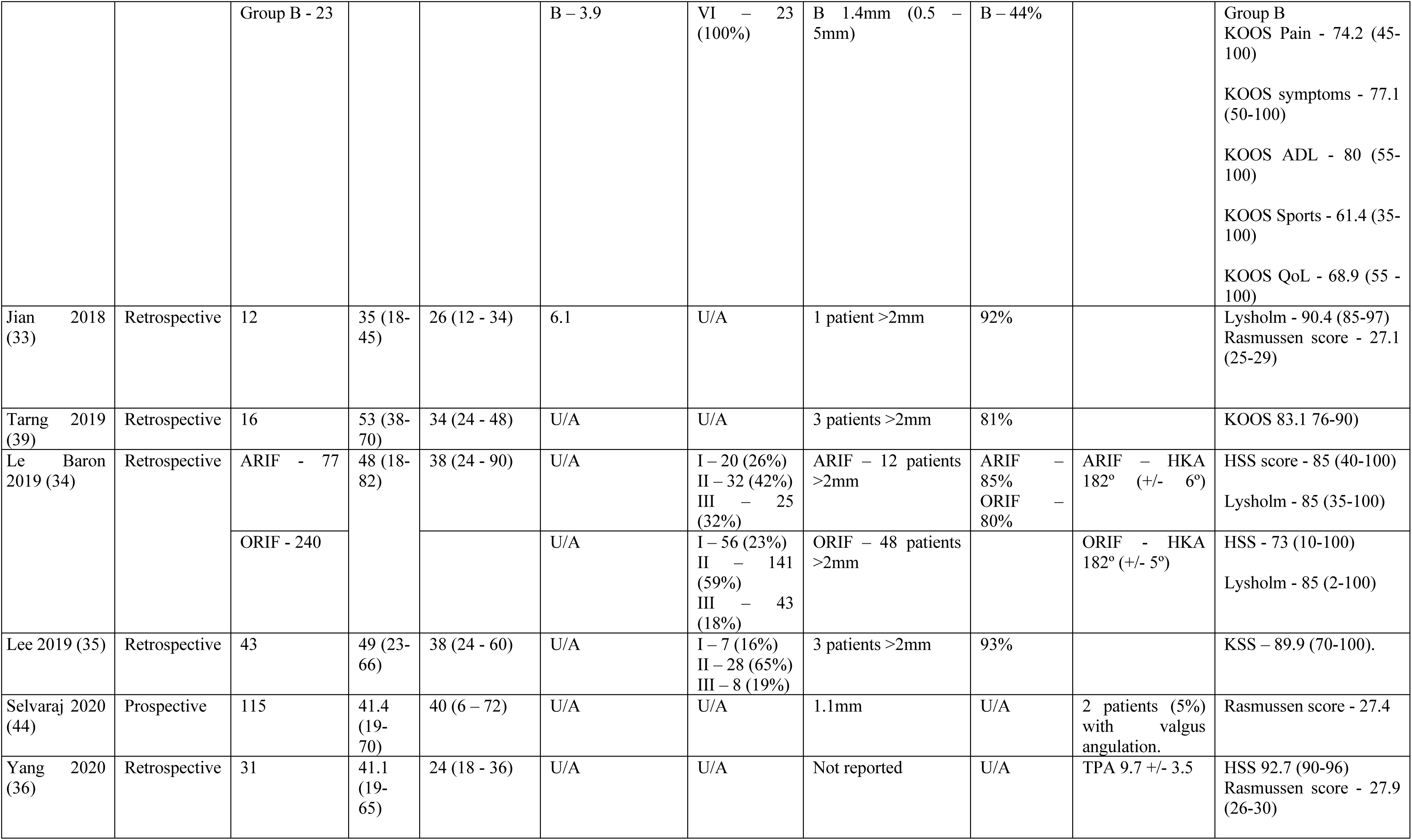
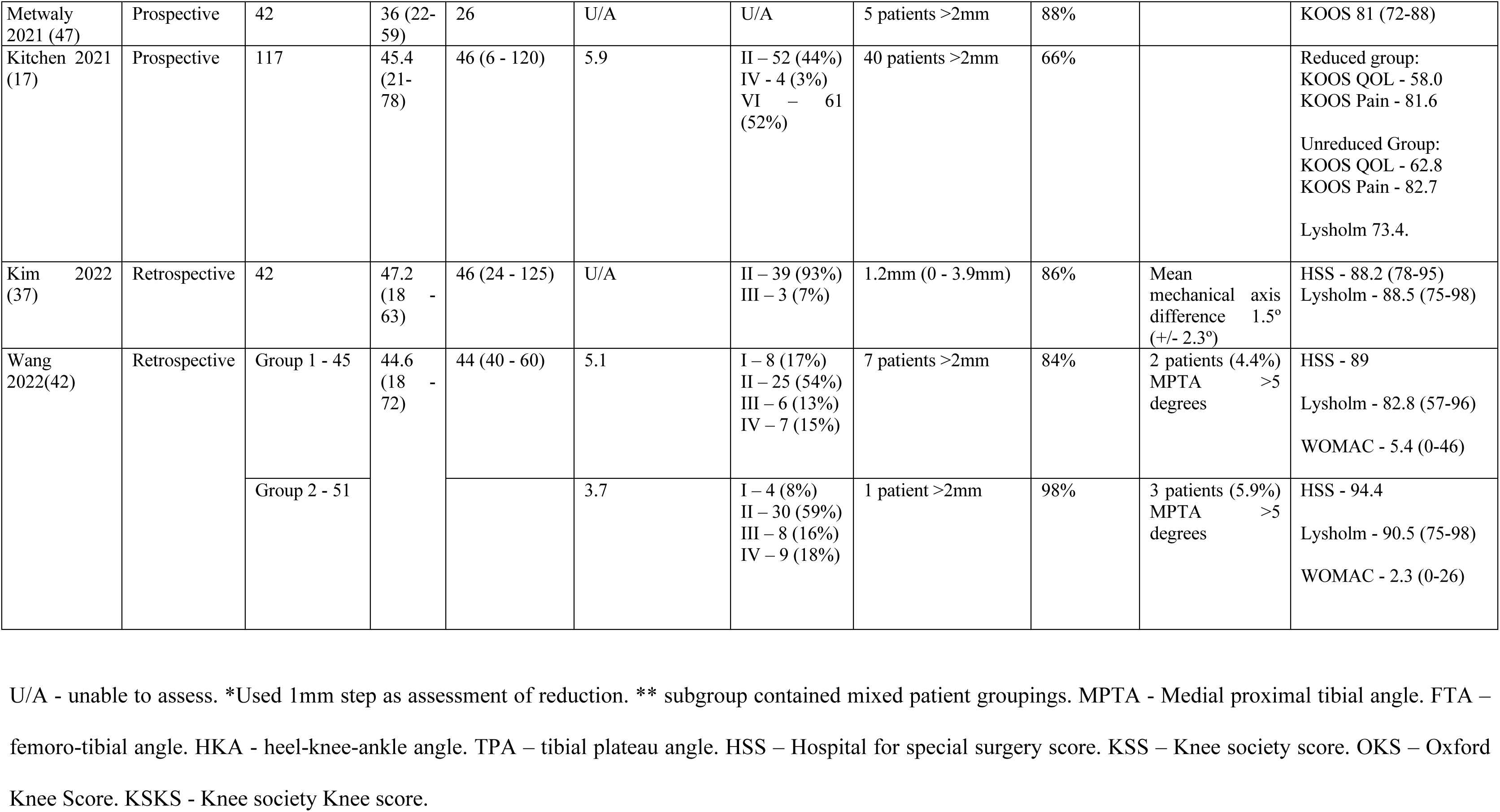
Key features of included studies.

### Study quality

**Table 2** shows the risk of bias assessment for the studies analysed using the RoBANS tool. Four studies were found to be of unclear risk of selection bias (7, 22–24), the remaining 26 studies were deemed to be low risk for selection bias. The low-risk studies reported on selection as either consecutive patients or all patients within a defined time-period meeting inclusion criteria.

**Table 2.** Risk of bias assessment for non-randomised studies.

| **Study** | **Selection of Participants** | **Confounding variables** | **Measurement of exposure** | **Blinding of outcome assessment** | **Incomplete outcome data** | **Selective reporting** |
| --- | --- | --- | --- | --- | --- | --- |
| Rossi 2008 (45) | Low risk | Low risk | Low risk | High risk | Low risk | Low risk |
| Chan 2008 (6) | Low risk | Unclear risk | Low risk | High risk | Low risk | Low risk |
| Levy 2008 (7) | Unclear risk | High risk | Low risk | High risk | Low risk | Low risk |
| Brunner 2009 (22) | Unclear risk | High risk | Low risk | Low risk | Low risk | Low risk |
| Luo 2010 (46) | Low risk | Low risk | Low risk | High risk | Low risk | Low risk |
| Siegler 2011 (25) | Low risk | High risk | Low risk | High risk | Unclear risk | Low risk |
| Ehlinger 2012 (26) | Low risk | High risk | Low risk | High risk | High risk | Low risk |
| Chiu 2013 (27) | Low risk | High risk | Low risk | Low risk | Low risk | Low risk |
| Persiani 2013 (23) | Unclear risk | Unclear risk | Low risk | High risk | Low risk | Low risk |
| Berkes 2014 (41) | Low risk | Unclear risk | Low risk | Low risk | Low risk | Low risk |
| Khatri 2014 (28) | Low risk | High risk | Low risk | High risk | Low risk | Low risk |
| Parkkinen 2014 (40) | Low risk | Unclear risk | Low risk | High risk | Low risk | Low risk |
| Ehlinger 2015 (38) | Low risk | High risk | Low risk | High risk | Low risk | Low risk |
| Lin 2015 (29) | Low risk | High risk | Low risk | High risk | Low risk | Low risk |
| van Dreumel 2015 (9) | Low risk | High risk | Low risk | High risk | Low risk | Low risk |
| Gavaskar 2016 (43) | Low risk | High risk | Low risk | Low risk | Low risk | Low risk |
| Elsøe 2016 (30) | Low risk | High risk | Low risk | High risk | Low risk | Low risk |
| Liu 2016 (24) | Unclear risk | High risk | Low risk | Low risk | Low risk | Low risk |
| Shimizu 2016 (31) | Low risk | High risk | Low risk | High risk | Low risk | Low risk |
| Ollivier 2018 (32) | Low risk | High risk | Low risk | High risk | Low risk | Low risk |
| Jian 2018 (33) | Low risk | High risk | Low risk | High risk | Low risk | Low risk |
| Tarng 2019 (39) | Low risk | High risk | Low risk | High risk | Low risk | Low risk |
| Le Baron 2019 (34) | Low risk | High risk | Low risk | Low risk | Low risk | Low risk |
| Lee 2019 (35) | Low risk | High risk | Low risk | High risk | Low risk | Low risk |
| Selvaraj 2020 (44) | Low risk | High risk | Low risk | High risk | Low risk | Low risk |
| Yang 2020 (36) | Low risk | High risk | Low risk | High risk | Low risk | Low risk |
| Metwaly 2021 (47) | Low risk | Unclear risk | Low risk | High risk | Low risk | Low risk |
| Kitchen 2021 (17) | Low risk | Low risk | Low risk | Low risk | Unclear risk | Low risk |
| Kim 2022 (37) | Low risk | High risk | Low risk | High risk | Low risk | Low risk |
| Wang 2022 (42) | Low risk | Low risk | Low risk | Low risk | Low risk | Low risk |

Eighteen (7, 9, 22, 25–39) of the 22 retrospective studies (82%) were deemed to be at high risk of bias to confounding variables due to limited data or lack of consideration for known variables in the study design or results. Three of the remaining four retrospective studies (23, 40, 41) were rated as unclear risk of confounding variables, as some of the known confounding factors affecting outcomes in patient demographics, such as smoking status, were taken into account, while other confounders were not recorded. Only one retrospective study (42) was rated as having a low risk of confounder bias, as the authors had reported on and balanced confounders in their study despite its retrospective nature. Comparatively, out of the eight prospective studies only two studies (25%) had an obvious high risk of bias (43, 44) with minimal consideration of confounders. Four were deemed as low risk for confounder bias (17, 24, 45, 46). The remaining two (6, 47) were deemed as having an unclear risk of bias, as some but not all confounders were assessed.

Eight of the studies (17, 22, 24, 27, 34, 41–43) explicitly stated that radiological or clinical outcome determination was performed by blinded assessors and were thus deemed at low risk of bias in this category (Table 2). The remaining studies did not report on blinding of assessors and were deemed as high risk of bias in this area. Two studies (17, 25) were deemed as having unclear risk of attrition bias due to incomplete outcome data being reported.

### Study Characteristics

Of the studies included, 22 (73%) were a retrospective design (7, 9, 22, 23, 25–42), with the remaining 8 being prospective (6, 7, 17, 24, 43–47). The mean follow-up for all studies was 40.0 (24 to 87) months. Of the included 30 studies, 9 (30%) were comparative studies assessing arthroscopic-assisted reduction with internal fixation (ARIF) in TPF (6, 7, 25, 27, 34, 36, 37, 45, 47). Four studies (13%) had a primary aim of comparing fixation techniques with outcomes (26, 33, 38, 42). Fifteen studies assessed fracture type based on the Schatzker classification system (6, 7, 9, 17, 26–28, 30, 31, 35, 37, 38, 42, 45, 46), with 729 TPFs classified this way. These consisted of 34 (4.7%) Schatzker I, 302 (41.4%) Schatzker II, 81 (11.1%) Schatzker III, 47 (6.4%) Schatzker IV, 80 (11.0%) Schatzker V and 185 (25.4%) Schatzker VI fractures.

### Articular reduction

Of the studies reviewed, 28/30 (93%) included specific reference to articular surface reduction. Two studies did not include any data on articular surface reduction but reported the Rasmussen radiological score and coronal alignment (25, 36). Twenty-four of the 30 of the studies reported the number of cases that had articular reduction to within 2 mm. 86% of cases within these 24 study cohorts reported an articular reduction rate of <2 mm. The ARIF group (9 studies) as a cohort had rate of successful reduction of 88.9% (75 - 100%), with 2 studies having 100% of cases with no articular step of greater than 2 mm (27, 45), and three studies in which the rate of successful reduction within the study could not be calculated (6, 7, 36). The percentage of a cohort having achieved articular reduction in patients without arthroscopic surgery was 84.6% (44 - 100%), with 3 studies having 100% of patients with articular step of less than 2 mm (24, 29, 43). Studies involving Schatzker types I, II, and III reported rates of successful reduction ranging from 60% to 100%, while those with Schatzker types IV, V, and VI had rates ranging from 44% to 100% (**Table 1**). Three studies in the ORIF group did not have sufficient data to calculate the rate of successful reduction (23, 31, 44).

### Coronal alignment

Eighteen studies reported on coronal alignment (6, 7, 25, 27, 28, 30–32, 34, 36–40, 43–46). Seven studies reported the use of weight-bearing x-rays (6, 25, 27, 32, 37, 40, 43, 45). Three studies (28, 32, 39) used the medial proximal tibial angle (MPTA), two studies using femoro-tibial angle (FTA) (43, 46), two used tibial-plateau angle (TPA) (36, 46) and a single study used the heel-knee angle (HKA) for alignment in the coronal plane (34). Eleven studies did not specify how alignment was assessed.

### Functional outcome scores

Various outcome scores were utilised within the assessed studies, the distribution of which is depicted in **Figure 2**. Eleven different functional outcome measures were used in the studies analysed. The most widely employed outcome measures were the Lysholm score in 14 studies (46.7%), the Hospital for Special Surgery score (HSS) in nine studies (30%), the Knee and Osteoarthritis Outcome Score (KOOS) in seven studies (23.3%) and the Rasmussen clinical outcome in seven studies (23.3%). While relationships between individual PROM scores and the corresponding degree of reduction did not achieve statistical significance, there was an observed trend that cohorts with a higher percentage of reduction reported a higher mean PROM, particularly for KOOS Pain (*p* = 0.136), Lysholm (*p* = 0.125) and HSS (*p* = 0.173). However, this did not translate to an effect of reduction on KOOS QOL (**Fig. 3**).

**Figure 2.**
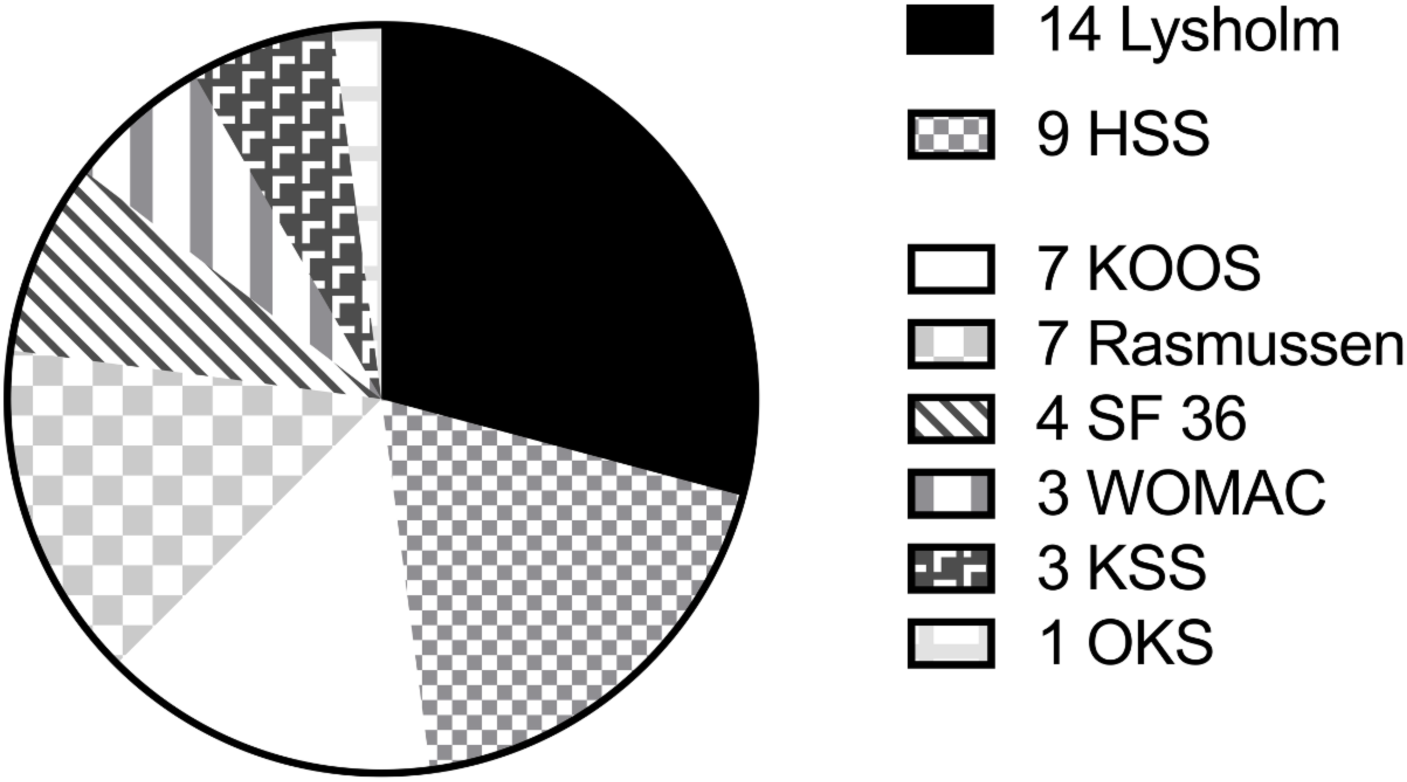
Distribution of functional outcome scores utilised between the included studies.

**Figure 3.**
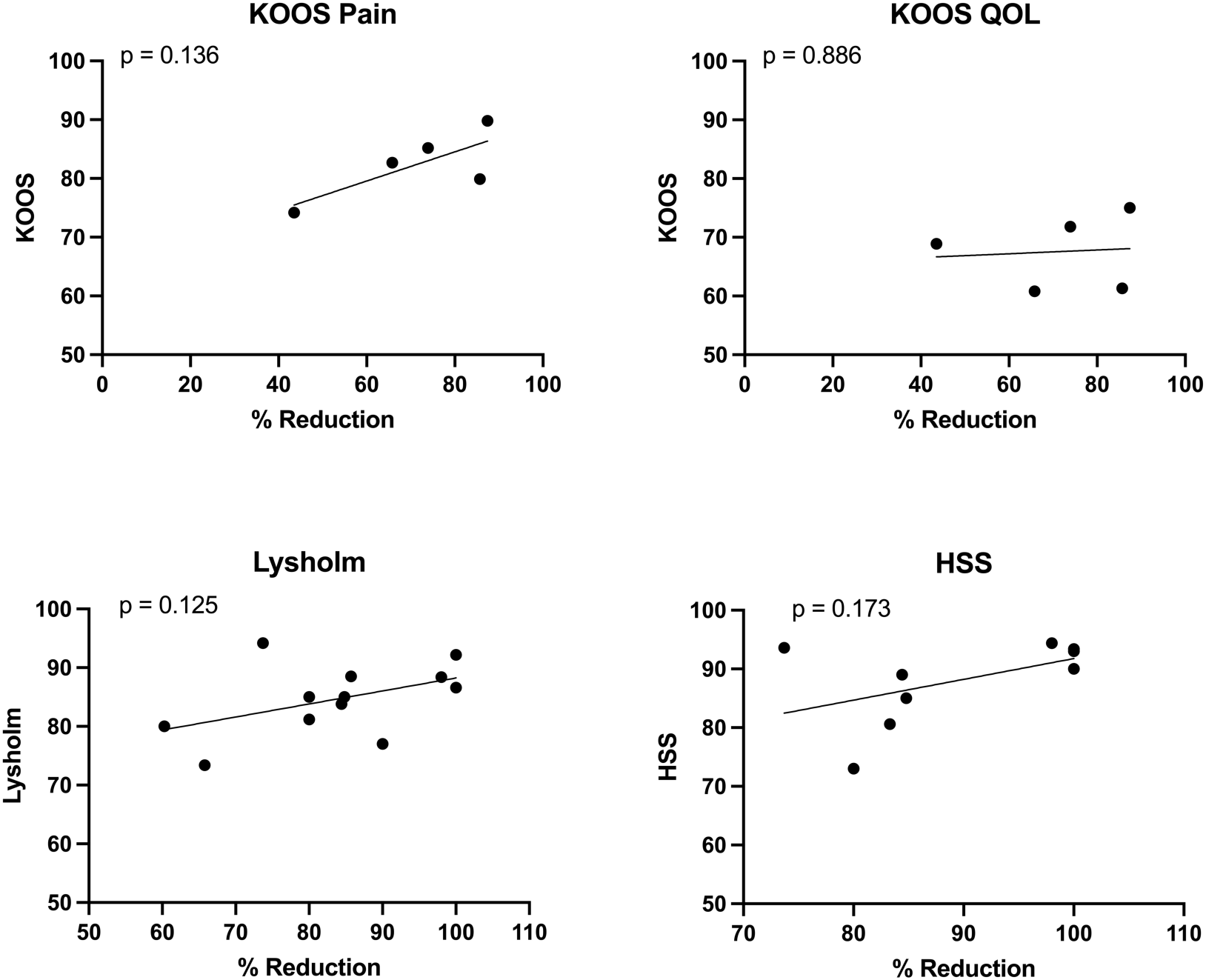
Rate of successful reduction vs PROM outcome score. HSS – Hospital for Special Surgery, KOOS – Knee Injury and Osteoarthritis Outcome Score, QOL – quality of life.

Lysholm, HSS, Rasmussen and KOOS (quality of life) have all previously had their outcome scores categorised into ‘good’ and ‘excellent’ scores (48, 49). Therefore, to account for the diversity of PROMs used, grouped analysis of studies, comparing good and excellent outcomes was performed (**Fig. 5**). On comparison of excellent vs good outcome scores and their percentage of cohort with articular reduction, a significant difference (*p* = 0.04) was found between the two groups. Those with excellent outcomes had a mean rate of successful reduction 10% higher than those with good outcomes.

Residual articular step of greater than 2 mm was associated with poorer outcome in some studies. Parkkinen *et al.* (40) demonstrated significant reduction in WOMAC scores (*p* = 0.02), as well as increased pain at night (*p* = 0.027), sitting (*p* = 0.013) and standing (*p* = 0.027) in those with articular depression of >2 mm (40). The results presented by Gavaskar *et al*. (43) demonstrated a statistically significant difference (*p* = 0.04) in the Lysholm score when anatomic (89 ± 4.4) and non-anatomic (83.3 ± 5.24) articular reductions were compared. Persiani *et al*. reported on mean articular reductions across several subgroups, all of which were greater than 2 mm, with four of the six subgroups having mean reductions of 4 mm or greater. Across these subgroups, the mean WOMAC scores were all 85 or greater out of a total possible score of 96, with higher scores being associated with worse outcomes. Interestingly, this contrasts with the Rasmussen clinical scores for the same subgroup of patients, all of which were greater than 20 and thus within the ‘good’ category, highlighting the differences between PROMS.

### Coronal Alignment

Regarding the relationship between coronal alignment and PROMs, a grouped analysis was not performed. Ehlinger *et al*. (38) reported that two patients within their study had mechanical axis deviation of 12 degrees each, with poor Lysholm (50 & 47) and HSS (58 & 45) scores when compared to the mean functional scores of the study of 80.1 and 83.3, respectively, although no statistical analysis was reported on this finding. Chan *et al*. reported two patients in their study with axial deviation of 10 degrees or greater (10 and 15 degrees) had fair functional outcomes using the Rasmussen scoring system (6). In contrast, Levy’s study (7) showed excellent Rasmussen scores in their small cohort of patients with moderate valgus alignments of three and seven degrees, with Rasmussen scores of 27 and 29 out of 30, respectively. Shimizu *et al*. (31) showed good clinical outcome with a Rasmussen score of 27, in their single patient with large valgus deformity (14 degrees). Tarng *et al*. (39) reported a mean MPTA of 84.68 ± 2.41 with a mean KOOS score 83.12 ± 5.01, with a single patient having five-degree malalignment with a KOOS score of 80. Le Baron *et al*. had two separate cohorts, one which underwent ARIF compared against an ORIF group, with comparable mean heel-knee angles (182 ± 6 & 182 ± 5, respectively) and Lysholm scores (85 ± 15.7 & 85 +/- 14.7, respectively) (34). Khatri *et al*. (28) reported a mean MPTA of 84.2 (range 83-92) resulting in a mean Oxford score of 42.6. Valgus alignment was shown to impact the development of post-traumatic osteoarthritis (OA) in those with more than 5 degrees of valgus alignment when using weightbearing mechanical axis on x-ray, with 6 of 12 (50%) developing OA when compared to only 1 of 20 (5%) with neutral coronal alignment (*p* = 0.006) (40).

## Discussion

The goal of surgery in TPF is to obtain and maintain articular reduction, restore joint alignment and biomechanics, while respecting the soft tissue envelope. This study demonstrates that cohorts in which >85% of cases achieved articular reduction within 2 mm reported significantly higher patient-reported outcome measures (PROMs) than those with lower reduction rates (see **Fig. 4**). Combining various PROMs such as Lysholm, HSS, Rasmussen, and KOOS into a subgroup analysis demonstrated that ‘excellent’ outcomes correlated with a higher percentage of a cohort being reduced compared to those achieving ‘good’ outcomes (*p* = 0.04). Individual studies reinforced this link, indicating that reductions of greater than 2 mm yield poorer outcomes compared to anatomical reductions, particularly when assessed using the WOMAC and Lysholm scores (40, 43).

**Figure 4.**
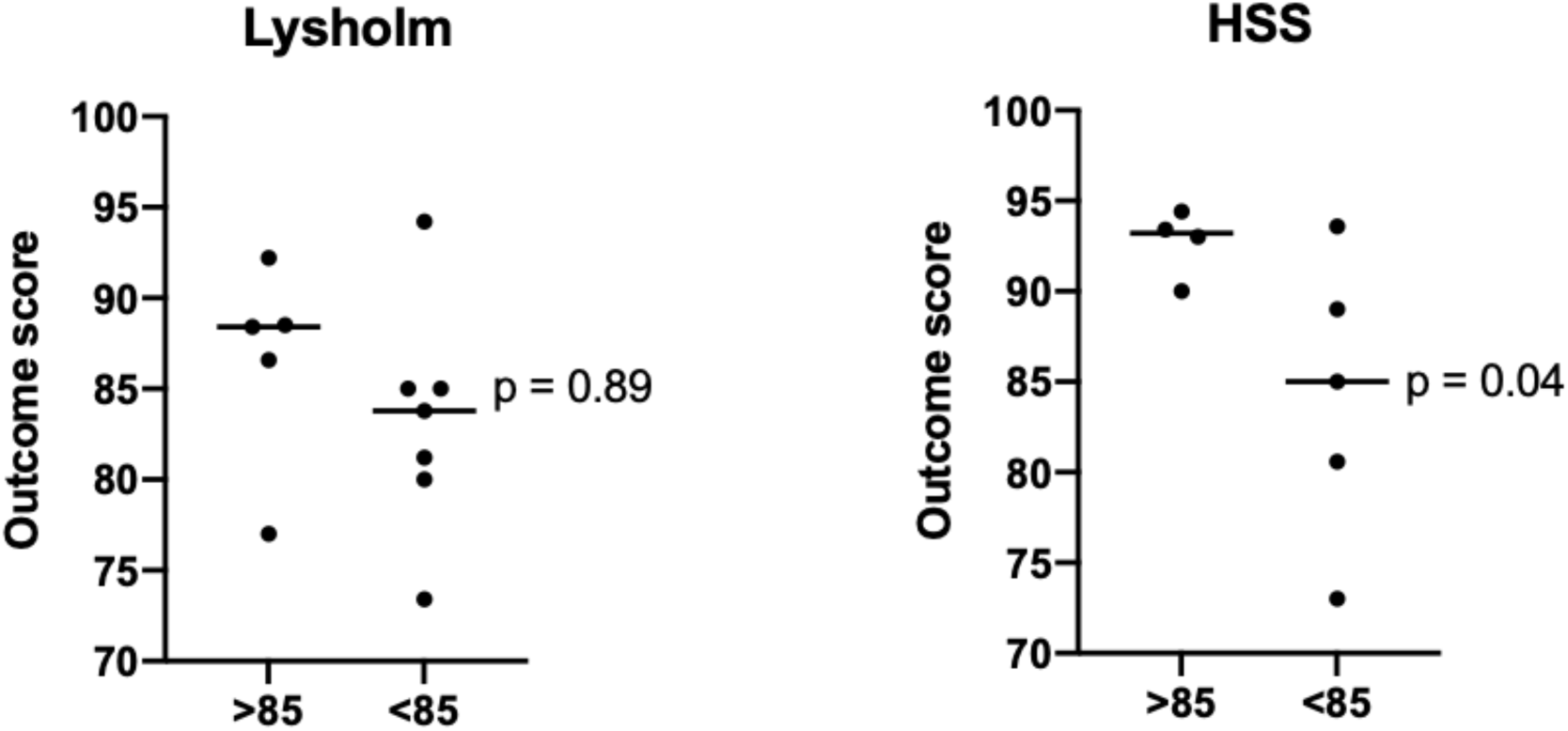
Grouped rate of successful reduction vs outcome score. Data was collated into two separate groups based on the mean articular reduction of 85% across all studies. A t-test was performed on each group to assess for difference in outcome score when rate of successful reduction was greater than or equal to 85% compared to less than 85% for Lysholm score and Hospital for Special Surgery Score (HSS).

**Figure 5.**
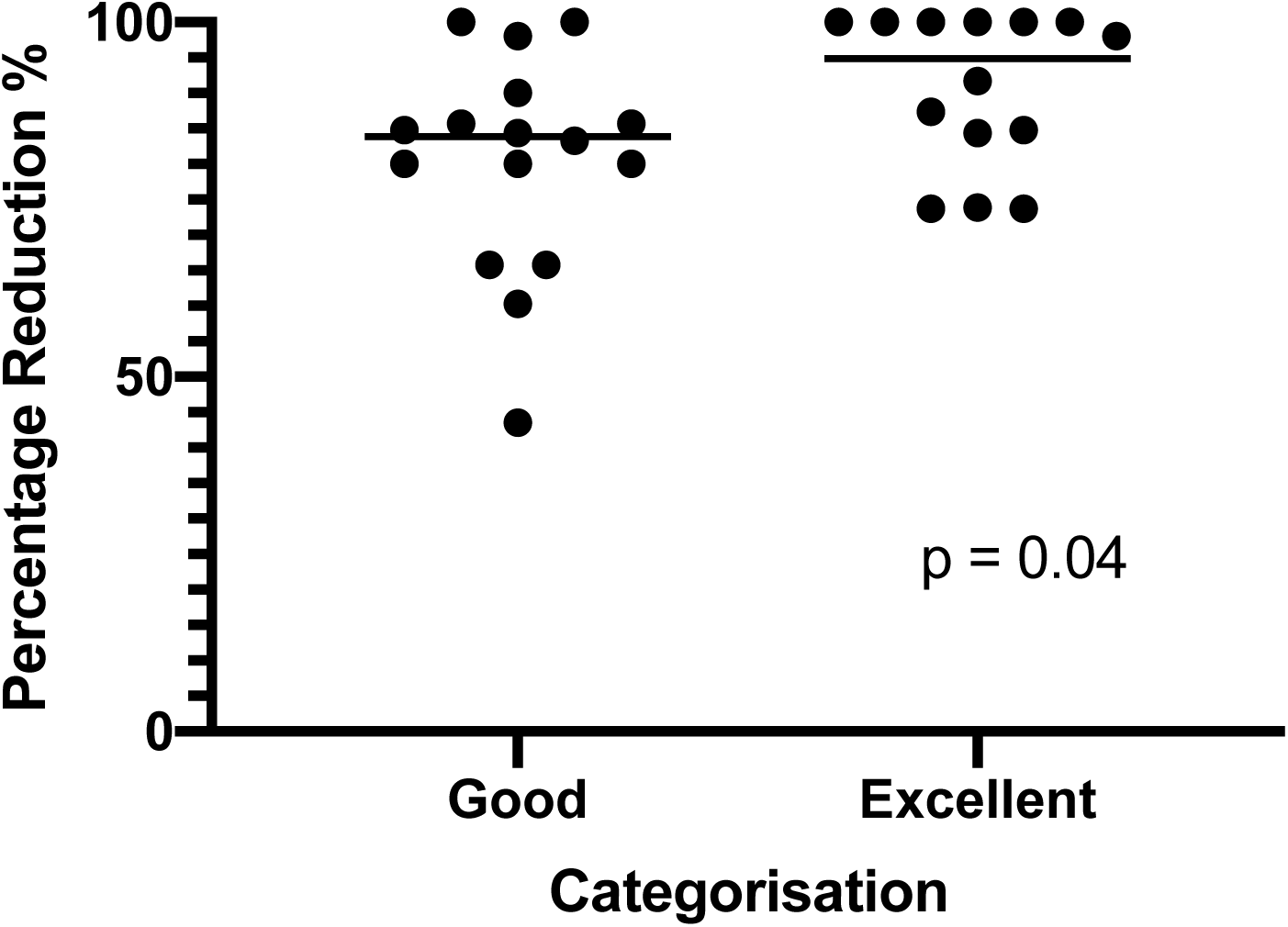
Rate of successful reduction when good and excellent outcome are achieved. Lysholm, HSS, Rasmussen and KOOS (quality of life) score that corresponded to good and excellent outcome scores were assessed against percentage articular reduction within studies using a t-test.

Although coronal malalignment is associated with degenerative changes in the knee (50), its impact on TPF outcomes is less definitive (16, 40, 51). Due to varied reporting on coronal alignment, a meta-analysis was not feasible in the current study. Some studies noted significant outcome reductions with malalignment exceeding 10 degrees of valgus (6, 38), while others reported favourable outcomes in cases with moderate deformities (3-7 degrees) (7) and even in severe valgus (14 degrees) (31). This supports recent findings that moderate malalignment may have limited impact on patient outcomes (51).

A challenge in evaluating TPF outcomes is the heterogeneity in PROMs and inconsistent definitions of acceptable articular reduction. Across the 30 studies assessed, 11 different PROMs were used, complicating collective analysis. The Lysholm score was the most frequently used (47%), followed by HSS (30%), KOOS (23%), and Rasmussen (23%). To address variability, outcome scores across Lysholm, HSS, KOOS quality of life (QOL), and Rasmussen were grouped, showing higher rates of successful reduction in cohorts with ‘excellent’ outcomes compared to ‘good’ outcomes (p = 0.04). This novel approach enhances statistical relevance and allows comparison across diverse measures. The KOOS QOL domain was chosen for its clarity in differentiating between bad, good, and excellent outcomes. Although the impact of pain on outcomes is significant, quality of life is considered a more holistic measure of patient satisfaction post-intervention (52). These findings suggest that precise reduction is crucial for improved outcomes.

One reason for the wide range of PROMs may be the lack of a universally accepted tool specifically for TPF. Existing PROMs, such as the Lysholm (developed for ACL surgery in 1982) (53), the HSS (for knee replacement in 1976) (54), and KOOS (for ACL and meniscal surgery in 1998), are not specifically designed for TPF. The only PROM originally intended for TPF, the Rasmussen score (13), is over 50 years old. The variability in PROM use not only complicates comparisons but also has implications for cost and patient participation (55). Over half of the studies (60%) reported more than one PROM, adding complexity to the interpretation of results. For instance, Persiani et al. (2013) found discrepancies in outcome in which high articular steps correlated with poor WOMAC scores but favourable Rasmussen outcomes (23), suggesting that some PROMs may lack the sensitivity needed for TPF-specific insights.

Additionally, the clinical significance of changes in PROM scores remains unclear due to the absence of defined minimum clinically important differences (MCID) (56) for most scores. The WOMAC score is an exception, with an MCID set at 6.7 points for TPF (55). Other PROMs, such as Lysholm and KOOS, lack MCID values for TPF, complicating the interpretation of outcomes (Ogura et al., 2021; Kitchen et al., 2021). Defining these thresholds would help ensure that reported changes are clinically relevant.

Limitations of the studies identified included the predominance of retrospective designs (73%), many of which showed a high risk of confounder bias (82%) (7, 9, 22, 23, 25–42). Only eight studies (27%) employed blinded outcome assessment, and just four (13%) were considered at low risk of confounder bias (17, 42, 45, 46). Additionally, publication bias may have influenced the overall high rate of successful reduction reported (85%), particularly with 30% of studies involving ARIF. However, in contrast to this, more than two-fifths (42.8%) of the studies involved high-grade Schatzker fractures (types IV, V, VI), which are complex and often yield worse outcomes.

In conclusion, evidence indicates that achieving higher rates of articular reduction are associated with improved PROMs, especially with the HSS score. In addition, patients attaining reductions to within 2 mm are more likely to report excellent outcomes than those with inadequate reduction. Future studies should focus on establishing clear MCID values for commonly used PROMs in TPF, with a particular emphasis on the Lysholm score, to enhance the validity and relevance of outcome assessments.

## Supporting information

Appendex A

## Data Availability

All data produced in the present study are available upon reasonable request to the authors

## Acknowledgements

This study received no specific funding.

