## Supplementary material for "Patients Report Better Outcomes Following Reduction of Tibial Plateau Fractures: A Systematic Review": Appendex A

Embase <1974 to 2023 May 5>
1 exp proximal tibia fracture/ 1825
2 exp proximal tibia/ and exp fracture/ 815
3 (((proximal* adj4 tibia*) or tibia*-plateau) and (angulat* or depress* or fracture*)).ti,ab,kf. 5448
4 1 or 2 or 3 6311
5 fracture fixation/ 25927
6 exp plate fixation/ 7987
7 exp osteosynthesis/ 46902
8 exp fracture reduction/ 13923
9 (fixat* or operat* or surg* or gleitosteosynthes* or osteo-synthes* or osteosynthes* or plate-stabili#ation? or bone-plate? or (fracture adj2 reduction?)).ti,ab,kf.
4265475
10 5 or 6 or 7 or 8 or 9 4281182
11 4 and 10 4436
12 limit 11 to conference abstracts 421
13 11 not 12 4015
14 limit 13 to (english language and yr="2000 -Current") 3068
15 (exp animal/ or exp animal experiment/) not ((exp animal/ or exp animal experiment/) and human/) 5529753
16 14 not 15 2853
